# Polygenic Risk for Depression is Associated with the Severity and Rate of Change in Depressive Symptoms Across Adolescence

**DOI:** 10.1101/2019.12.31.19016212

**Authors:** Alex S. F. Kwong, Tim T. Morris, Rebecca M. Pearson, Nicholas J. Timpson, Frances Rice, Evie Stergiakouli, Kate Tilling

## Abstract

Adolescence marks a period where depression will commonly onset and previous research using twin studies has suggested that genetic influences play a role in how depression develops and changes across adolescence. Recent genome-wide association studies have also shown that common genetic variants – which can be combined into a polygenic risk score (PRS) – are also implicated in depression. However, the role of PRS in adolescent depression and changes in adolescent depression is not yet understood. We aimed to examine the association between a PRS for depressive symptoms and depressive symptoms across adolescence and young adulthood, and how polygenic risk is associated with changes in depressive symptoms using two methods: cross-sectional analysis and multilevel growth curve modelling to examine the rate of change over time. Using data from over 6000 participants of the Avon Longitudinal Study of Parents and Children (ALSPAC) we examined associations between genetic liability to depressive symptoms (PRS for depressive symptoms) and self-reported depressive symptoms (short mood and feelings questionnaire over 9 occasions from 10-24 years). We examined cross-sectional associations at each age and longitudinal trajectories of depressive symptoms in a repeated measures framework using growth curve analysis. The PRS was associated with depressive symptoms throughout adolescence and young adulthood in cross-sectional and growth curve analyses, though associations were stronger in the latter analyses. Growth curve analyses also provided additional insights, demonstrating that individuals with a higher PRS had steeper trajectories of depressive symptoms across adolescence with a greater increasing rate of change. These results show that common genetics variants as indexed by a PRS for depressive symptoms influence both the severity and rate of change in adolescent depressive symptoms. Longitudinal data that make use of repeated measures designs have the potential to provide greater insights into the factors that influence the onset and persistence of adolescent depression.

## Introduction

Depression is a common mental health disorder and predicted to be the highest global burden of disease by 2030 [1]. Adolescence marks a period where depressive symptoms increase and major depressive disorder will commonly onset [2-5]. Adolescent depressive symptoms and major depressive disorder are associated with a number of psychiatric and social impairments in later life and show strong continuity with depression in adult life, thus making it important to prevent and treat [6-9].

Depression has a complex and multifactorial aetiology, comprised of both environmental and genetic contributions [10, 11]. Adult twin studies have estimated that the heritability of major depressive disorder is between 31% - 42% [12]. Twin studies of depressive symptoms during adolescence have estimated similar heritability to that of adult depression ~ 40% [13] with lower estimates reported for symptoms during childhood. There is considerable variability in heritability estimates between different studies depending on age, informant and measure of assessment [14]. Depressive symptomatology typically increases during adolescence and twin studies have shown that genetic contributions influence this developmental change [15]. In particular, genetic contributions may increase throughout development [16, 17] although some inconsistent results have also been reported [18]. There are strong continuities reported between adolescent and adult depression [9], so the inconsistent estimates of heritability for adolescent depressive symptomatology are puzzling and may be due to between-study differences in measurement and age of the sample studied. Longitudinal data spanning early adolescence and young adulthood using the same assessments and respondents over time would aid understanding of the nature of genetic influences on onset and persistence of symptoms across development.

Recent advances in genome-wide association studies (GWAS) have provided evidence that common genetic variation plays a role in depression [19–21] with many genetic variants or single nucleotide polymorphisms (SNPs) each having a small effect [22]. Polygenic risk scores (PRS), which sum the number of “risk” variants that an individual possesses for a trait weighted by their effect size [23], can be used as an indicator of an individual’s genetic liability to depression. Several studies have used depression PRS taken from GWAS of major depressive disorder or depressive symptoms in adult populations to investigate how they influence depressive symptomatology over development in younger populations [24-27]. One study found that a PRS for major depression was associated with depression in a clinical and population cohort of children and adolescents [26], whilst another found that the influence of a depression PRS on emotional problems increased with age with weaker effects in childhood which developed in adulthood [27]. Two other studies have since used a developmental trajectory approach to identify ‘classes’ of individuals whose depressive symptoms differ across adolescence [24, 25]. These studies found evidence of differing trajectory classes based on age of onset and persistence of symptoms over time that were associated with depression PRS. Together these studies highlight that polygenic risk is likely to play a role in the development and maintenance of adolescent depression. However, these studies do not tell us how polygenic risk for depression may vary by age, more that polygenic risk is associated with a typology of depression (i.e., adolescent onset depression or those with persistent symptoms). Further work examining how polygenic risk for depression varies by ages could help determine when genetic liability for depression may be affecting changes in depressive symptoms at important stages of development. This in turn is necessary for uncovering the mechanisms underlying depression.

There is evidence that PRS are associated with changes across childhood and adolescence for other traits such as height [28] and BMI [29, 30]. These studies have all used a repeated measures framework (i.e., to estimate trajectories or growth curve models) to examine genetic influences for changes in a trait. Work suggests that using a repeated measures framework such as growth curve modelling may help improve the statistical power of genetic analysis [31]. For example, measurement error and low statistical power are problems in genomic analysis as genetic effects tend to be small in magnitude and require large sample sizes with precision to detect true effects [32]. Likewise, variation in the reported genetic component for depression (i.e., heritability) may be partially a result of differential measurement error at different assessment occasions [13]. However, a longitudinal approach which uses repeated measures may reduce measurement error and noise by increasing statistical power as there are multiple occasions included in the analysis, rather than just one occasion [33]. Multiple measurements also maximise the number of participant responses and may obtain a more precise estimate of an individual’s “true” latent trait score as the assessment is repeated over time, and not just at one occasion. Using these repeated measurements it is also possible to reduce the burden for multiple testing that would occur when looking at associations across timings in a growth curve setting as the number of multiple comparisons are reduced [34]. Repeated measures analysis, in particular growth curve modelling, may provide an advantage to traditional cross-sectional analysis and also quantify how and when a trait changes over time, which in this context could help further explain the role of genetics in changes to adolescent depression over time.

The aim of this study was to examine how genetic liability for depression (as indexed by a PRS for depressive symptoms) influenced depressive symptoms across adolescence and early adult life using cross-sectional and repeated measures designs. Specifically, we aimed to test how a depressive symptoms PRS was associated with both the initial level and the rate of change of depressive symptoms over this developmental risk period for depression. We conducted the following analyses: 1) we used a PRS taken from a recent GWAS of depressive symptoms [35], and examined associations at nine separate occasions in a UK based population cohort between the ages of 10 and 24 years old (cross-sectional analysis). 2) we then used growth curve modelling to construct trajectories of depressive symptoms in the same cohort and examined how the PRS for depressive symptoms was associated with the rate of change in depressive symptoms throughout adolescent development. 3) finally, we examined if a higher PRS was associated with differences in depressive symptoms scores across this developmental period, in order to determine when the PRS would be having its greatest effect of depressive symptomology.

## Methods

### Sample

We used data from the Avon Longitudinal Study of Parents and Children (ALSPAC), a longitudinal cohort study that recruited pregnant women residing in the former area of Avon, UK with expected dates of delivery 1st April 1991 to 31st December 1992 [36, 37]. The initial cohort consisted of 14,062 live births, but has been increased to 14,901 children who were alive after one year with further recruitment [38]. Ethical approval was obtained from the ALSPAC Ethics and Law Committee and the Local Research Ethics Committees. The study website contains details of all the data that is available through a fully searchable data dictionary and variable search tool: http://www.bristol.ac.uk/alspac/researchers/our-data.

### Depressive Symptoms

Self-reported depressive symptoms were measured on nine occasions between ages 10 and 24 using the short mood and feelings questionnaire (SMFQ) [39]. See Table 1 for the ages at which the SMFQ was assessed. The SMFQ is a 13-item questionnaire that measures the presence of depression symptoms in the last two weeks and was administered via postal questionnaire or in research clinics. Each item is scored between 0-2, resulting in a summed score between 0-26. The SMFQ correlates highly (*r* = 0.58) with clinical depression [40, 41].

**Table 1.** Descriptive statistics of the Short Mood and Feelings Questionnaire (SMFQ) for individuals included in this analysis

| Occasion<br>(Total *N*) | Mean Age | Mean SMFQ | Alpha | % Above SMFQ<br>Threshold* |
| --- | --- | --- | --- | --- |
| 1 (*N* = 5,320) | 10.64 (0.25) | 4.00 (3.49) | .80 | 5.85 |
| 2 (*N* = 4,928) | 12.81 (0.23) | 3.94 (3.84) | .84 | 7.11 |
| 3 (*N* = 4,495) | 13.83 (0.21) | 4.92 (4.49) | .86 | 11.81 |
| 4 (*N* = 3,524) | 16.68 (0.24) | 5.85 (5.63) | .91 | 17.68 |
| 5 (*N* = 3,212) | 17.82 (0.37) | 6.50 (5.21) | .90 | 21.05 |
| 6 (*N* = 2,389) | 18.64 (0.49) | 6.73 (5.85) | .91 | 21.20 |
| 7 (*N* = 2,380) | 21.95 (0.52) | 5.56 (5.46) | .91 | 17.44 |
| 8 (*N* = 2,707) | 22.88 (0.51) | 6.07 (5.40) | .90 | 17.64 |
| 9 (*N* = 2,737) | 23.86 (0.51) | 6.84 (5.91) | .91 | 23.49 |

Standard deviations are given in (parenthesis). \*Scores equal to or above 11 have been proposed as good indicators of depression [41]. Individuals included in this analysis had data on the SMFQ, genetic data to make the PRS/principal components of ancestry and data on sex. Alpha scores were created using Cronbach's Alpha.

### Polygenic Risk Score for Depressive Symptoms

The PRS for depressive symptoms was created in PRSice [42], using summary statistics from a recent genome wide association study (GWAS) of depressive symptoms on 161,460 individuals [35]. The PRS was created by weighting the effect sizes of up to 120,422 single nucleotide polymorphisms (SNPs) associated with depression symptoms from the initial GWAS at eight *P*-value thresholds (as 5×10^-08^, 5×10^-07^, 5×10^-06^, 5×10^-05^, 5×10^-04^, 5×10^-03^, 5×10^-02^ and 5×10^-01^). The PRS was standardised to have a mean of 0 and a standard deviation of 1, thus a higher PRS represents higher liability to depression symptoms. We included SNPs that had a MAF of > 1% and info score > 80%) and excluded SNPs with an R^2^ of >0.1, if they were within 250Kb of each other. We excluded SNPs located in the extended MHC region (chromosome 6 (26-33Mb)). Further genotyping information is available in the Supplement.

### Statistical Analysis

For the cross-sectional analysis, linear regression analysis was used to examine the association between depressive symptoms and the PRS at each of the nine occasions. *P* values were corrected for false discovery rate (FDR) due to the number of statistical tests (eight PRS thresholds x nine depressive symptoms occasions [72 tests]). Bootstrapping with 1,000 iterations was used to calculate confidence intervals for R^2^ (the amount of variance explained by the PRS).

For the repeated measures analysis, trajectories of depressive symptoms were estimated using multilevel growth-curve modelling [43, 44]. Briefly, multilevel growth-curve models create population averaged trajectories with intercept and slope terms. Individual level trajectories then vary around this population average (i.e., each person can have their own trajectory, with their own intercept and slope that deviates from the population average). Previous analysis of these data has shown that changes in depressive symptoms over time are non-linear [45, 46], with depressive symptoms rising until the age of about 18, then decreasing until around the age of 22, before rising again towards the age of 24. To model these non-linear trajectories, a multilevel quartic growth-curve polynomial model was chosen. This model contains five key parameters: the intercept, the linear age term, the quadratic age term, the cubic age term and the quartic age term. These age terms allow for non-linearity in the trajectory and changes in depressive symptoms. Previous research using this data to estimate multilevel growth curves has found that a quartic polynomial model best fitted the data [47]. We further assessed the feasibility of this model using information criteria and likelihood ratio tests, consistent with other studies using multilevel growth-curve models [48] – see Supplementary Tables S1 and S2, and Figure S2.

To examine how the PRS was associated with changes in the growth curve model, we included a main effect of the standardised PRS and an interaction of the PRS with each of the fixed-effects age polynomial terms (i.e., the linear, quadratic, cubic and quartic age terms). Age was grand-mean centred to 16.53 years (the mean age of all assessments) in order to improve interpretation, since model intercept and intercept variance then broadly correspond to the middle of adolescence [49]. The intercept and four polynomial age terms were allowed to vary randomly across individuals to capture each individual’s unique trajectory (i.e., a random intercept and random slopes model). Further information regarding model fit and the model equations can be found in the Supplement.

To assess the association between the depressive symptoms PRS and development of symptoms over time, we created one trajectory associated with low genetic liability (1 SD below mean) and one associated with low genetic liability (1 SD above the mean). We then calculated the predicted depressive symptoms scores at each of the following ages: 10.64, 12.81, 13.83, 16.68, 17.82, 18.64, 21.95, 22.88 and 23.86 (to coincide with the mean ages at which the SMFQ was assessed at each of the nine occasions), for both the greater and lower PRS trajectories. We compared the predicted depressive symptoms scores at each of these ages between greater and lower PRS trajectories. Further information on how these were calculated for the trajectories is presented elsewhere [47], but briefly the depressive symptoms scores are calculated at each age for the two trajectories (i.e., depressive symptom scores at age 12 for the low PRS and high PRS trajectory). Then using the delta method (which incorporates the estimate, standard errors and confidence intervals), these two scores are then compared to reveal an estimated difference that has measures of certainty and precision. Stata code for our analysis can be found here: https://github.com/kwongsiufung.

All analyses were conducted using Stata 15 (StataCorp, College Station, TX, USA), with trajectories analysis using the user-written runmlwin command [50], which calls the standalone multilevel modelling package MLwiN v3.01 (www.cmm.bristol.ac.uk/MLwiN/index.shtml). All analyses were adjusted for sex, age (only in the cross-sectional analyses, as the longitudinal analyses adjust for age by default) and the first ten principal components of ancestry.

### Missing Data

Missing data in the trajectories analysis were handled using full information maximum likelihood estimation (FIML) [51]. Briefly, this assumes that the probability of an individual missing a measure of depressive symptoms does not depend on their underlying depressive symptoms score at that occasion, given their observed depressive symptoms trajectory at other occasions. We included individuals into our analysis if they had at least one measurement of depression symptoms in order to maximise power [52]. Previous research on these data has shown that trajectory shapes and characteristics do not vary when comparing individuals with at least one or at least 4 measurements of depressive symptoms [45].

### Sensitivity Analyses

We conducted a variety of sensitivity analyses including (1) negative control analysis with a PRS for height and then the PRS for depressive symptoms on height trajectories, (2) additional sensitivity with a PRS taken from a recent GWAS of major depression, (3) tests for measurement invariance for depressive symptoms and (4) analysis exploring the association between the PRS and missing data patterns. Further information regarding these sensitivity analyses can be found in the Supplement.

## Results

### Sample Characteristics

Of the original 14,901 children alive after one year, 9,399 had at least one measurement of depressive symptoms and 7,877 had genotype data. For the cross-sectional analysis, data were available for 5,320 individuals with a measurement of depressive symptoms at age 10 and genotype data. However, sample size decreased to 2,737 individuals with both a depression symptoms measurement at age 24 and genotype data. Descriptive information can be found in Table 1 and a STROBE diagram along with further information is given in Supplementary Figure S1 and Table S3. For the repeated measures analysis, data were available for 6,305 individuals with at least one measurement of depressive symptoms and genotype data.

### Cross-sectional Association Between Depressive Symptoms Polygenic Risk Score and Depressive Symptoms

Given that more liberal PRS *P*-value thresholds have been used when examining the predictive capabilities of PRS for psychiatric traits [19], we focused on the more liberal PRS thresholds of 0.005, 0.05 and 0.5. For completeness, full estimates are given for analyses at all thresholds can be found in Supplement Tables S4-S13.

The PRS with a *P*-value threshold of 0.005 showed the strongest associations (*β* = 0.33) with depressive symptoms on average across occasions (Supplementary Tables S4-S13). Effect sizes tended to increase throughout adolescence and into young adulthood with the greatest association between a one standard deviation higher PRS and depressive symptoms observed at age 24 (*β* = 0.610, 95 CIs = 0.392, 0.828, *P* = 3.34×10^-06^), as shown in Figure 1 and Table 2. Additional analyses across the three most liberal thresholds (0.005, 0.05 and 0.05) yielded similar results and can be found in Supplementary Figure S5 and Supplementary Table S13.

**Figure 1.**
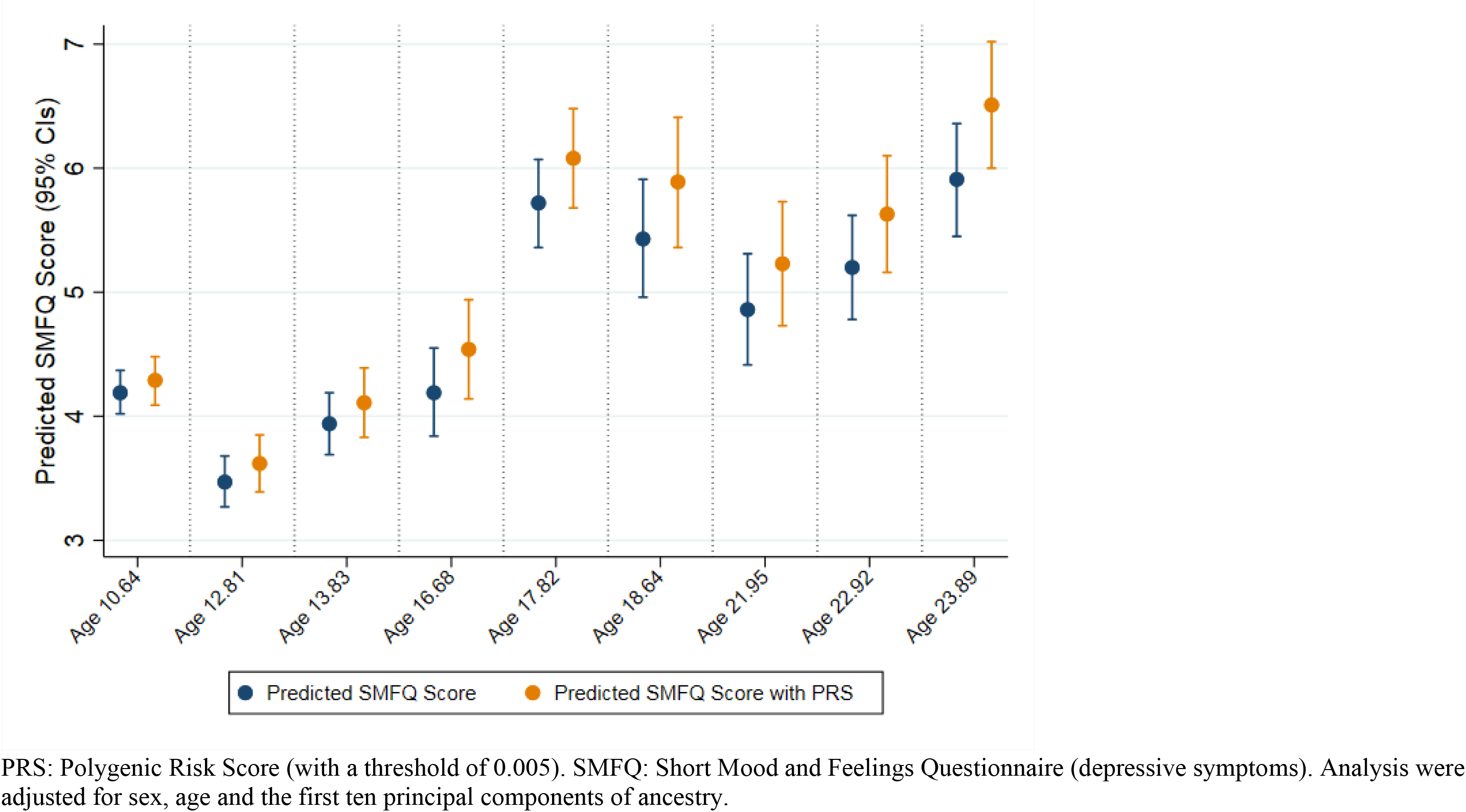
Cross-sectional analysis between the depressive symptoms PRS and depressive symptoms across adolescence with predicted SMFQ scores

**Table 2.**
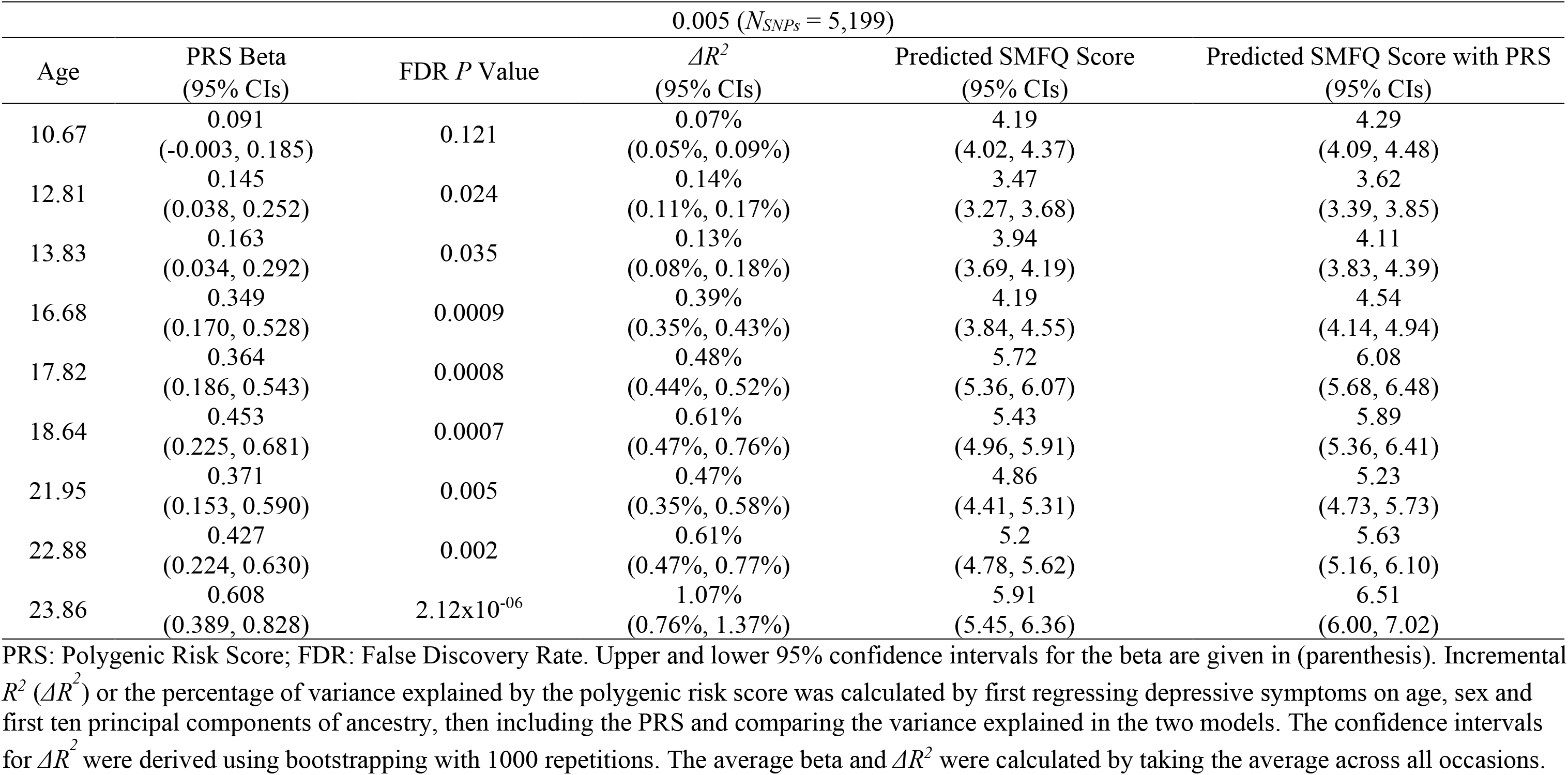
Association between the depressive symptoms PRS and depressive symptoms at various across adolescence

### Longitudinal Association Between Depressive Symptoms Polygenic Risk Score and Trajectories of Depressive Symptoms

We selected the PRS with a *P*-value threshold of .005 for the trajectory analysis because this PRS threshold showed stronger average effect sizes across the cross-sectional analyses and is a similar threshold to previous studies (see Table S13). Sensitivity analyses were conducted at other thresholds and showed almost identical results and inferences (see Supplementary Tables S14-S16).

A one standard deviation higher depressive symptoms PRS was associated with higher depressive symptoms at the intercept age of 16.53 (*β* = 0.363, 95 CIs = 0.230, 0.496, *P* = 8.56×10^-08^). The depressive symptoms PRS also showed evidence for change over time, with a one standard deviation higher PRS strongly associated with a linear change in depressive symptoms (*β* = 0.048, 95 CIs = 0.016, 0.080, *P* = 0.003). However, a one standard deviation higher PRS showed weak and inconsistent associations with the quadratic, cubic or quartic age terms (see Figure 2 and Table S13).

**Figure 2.**
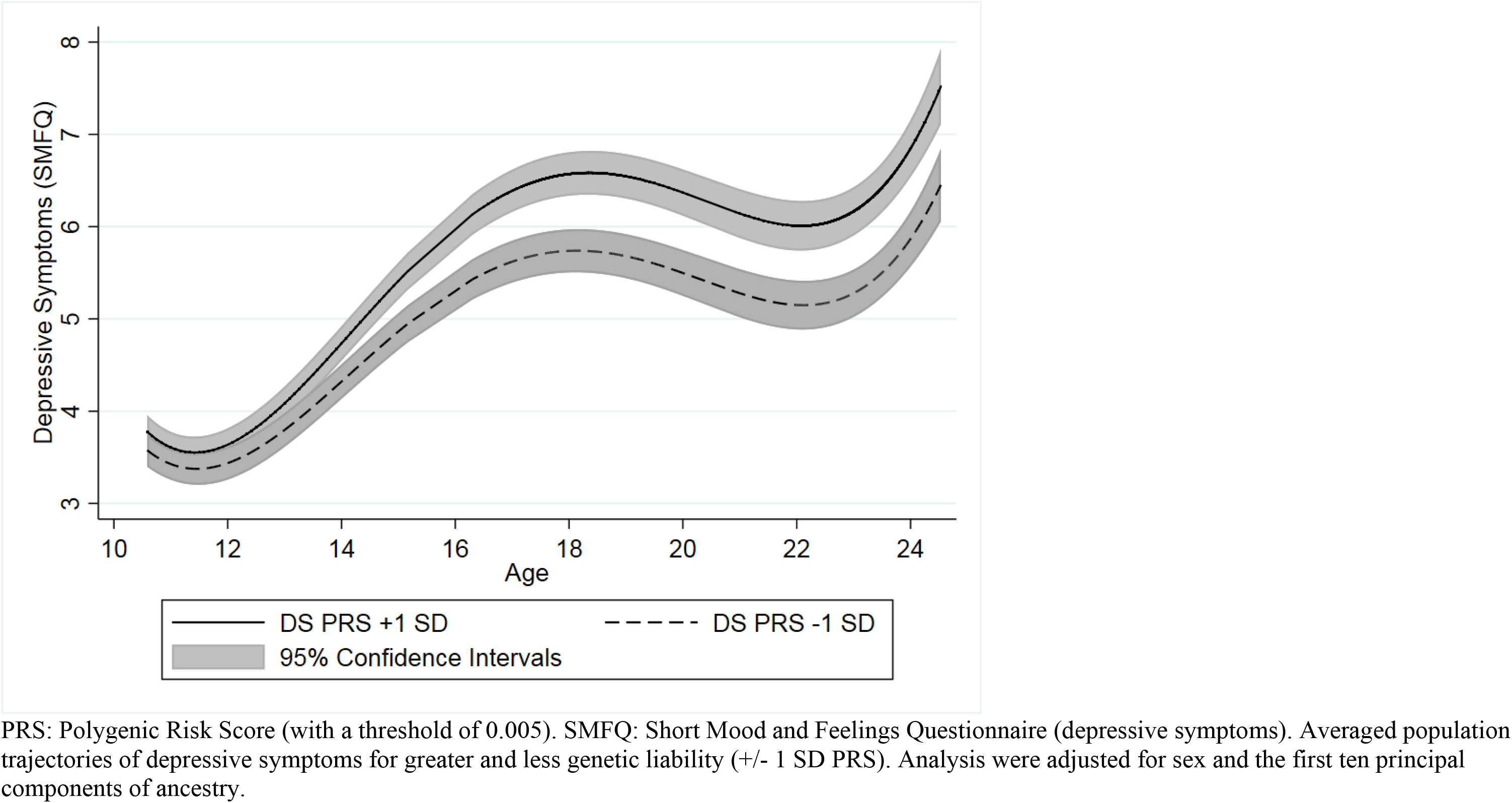
Association between depressive symptoms PRS and trajectories of depressive symptoms

### Comparisons Between Higher and Lower Genetic Risk at Various Ages

Our comparisons between those with higher risk (1 SD above mean in the PRS) and those with lower risk (1 SD below mean in the PRS) demonstrated strong evidence that depressive symptoms were higher for those with greater PRS risk at all ages of adolescence and young adulthood from age 12 onwards (*P*^FDR^*s* ≤ 0.037), and weaker evidence at age 10 (*P*^FDR^ = 0.063) (Table 3). The largest difference between these two trajectories was observed at age 24 with roughly a one-point difference of the SMFQ (*β*_diff_ = 0.980, 95 CIs = 0.580, 1.380, *P*^FDR^ = 1.24×10^-06^).

**Table 3.**
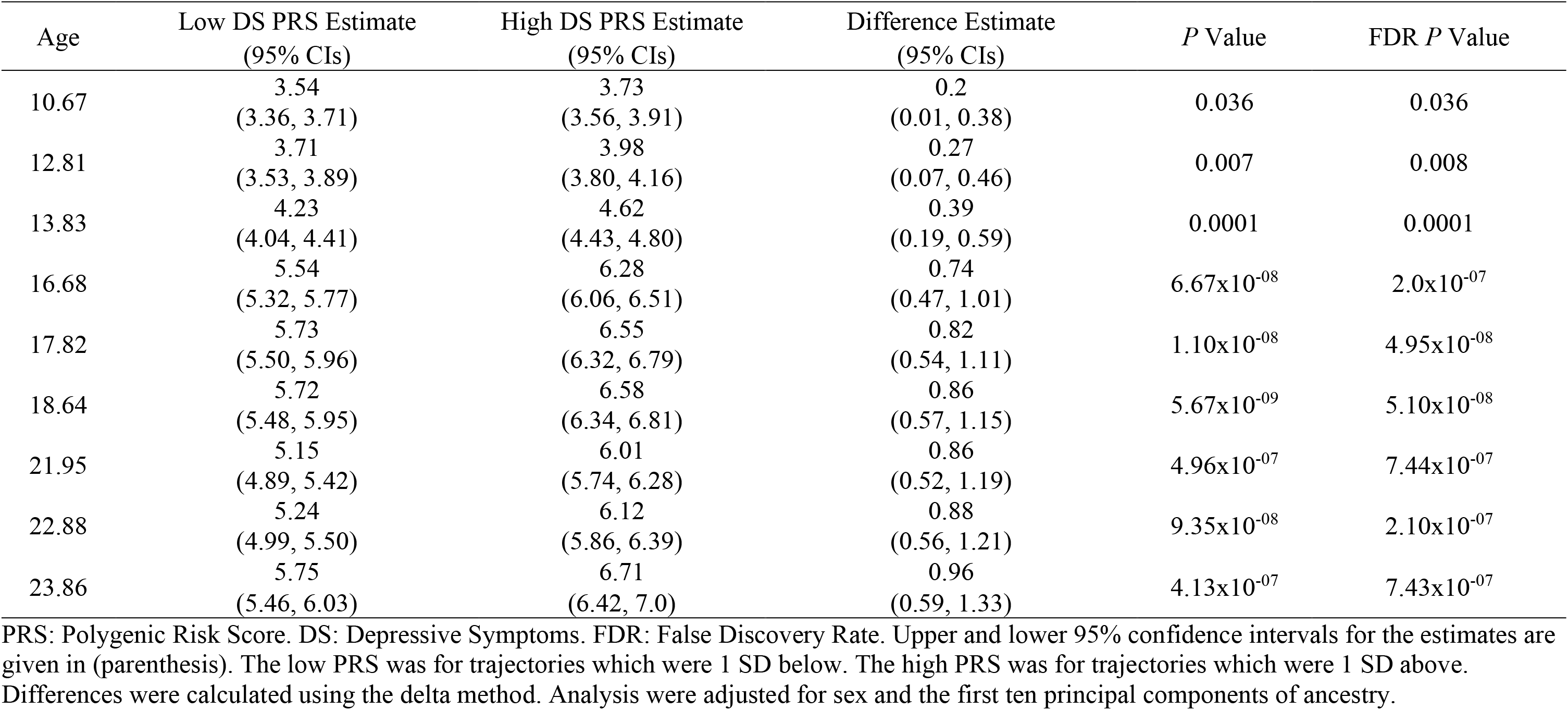
Comparisons across multiple ages between trajectories that were +/- 1 SD Above for the Depressive Symptoms PRS (*N*=6,305)

| Age | Low DS PRS Estimate<br>(95% CIs) | High DS PRS Estimate<br>(95% CIs) | Difference Estimate<br>(95% CIs) | *P* Value | FDR *P* Value |
| --- | --- | --- | --- | --- | --- |
| 10.67 | 3.54<br>(3.36, 3.71) | 3.73<br>(3.56, 3.91) | 0.2<br>(0.01, 0.38) | 0.036 | 0.036 |
| 12.81 | 3.71<br>(3.53, 3.89) | 3.98<br>(3.80, 4.16) | 0.27<br>(0.07, 0.46) | 0.007 | 0.008 |
| 13.83 | 4.23<br>(4.04, 4.41) | 4.62<br>(4.43, 4.80) | 0.39<br>(0.19, 0.59) | 0.0001 | 0.0001 |
| 16.68 | 5.54<br>(5.32, 5.77) | 6.28<br>(6.06, 6.51) | 0.74<br>(0.47, 1.01) | $6.67 \times 10^{-8}$ | $2.0 \times 10^{-7}$ |
| 17.82 | 5.73<br>(5.50, 5.96) | 6.55<br>(6.32, 6.79) | 0.82<br>(0.54, 1.11) | $1.10 \times 10^{-8}$ | $4.95 \times 10^{-8}$ |
| 18.64 | 5.72<br>(5.48, 5.95) | 6.58<br>(6.34, 6.81) | 0.86<br>(0.57, 1.15) | $5.67 \times 10^{-9}$ | $5.10 \times 10^{-8}$ |
| 21.95 | 5.15<br>(4.89, 5.42) | 6.01<br>(5.74, 6.28) | 0.86<br>(0.52, 1.19) | $4.96 \times 10^{-7}$ | $7.44 \times 10^{-7}$ |
| 22.88 | 5.24<br>(4.99, 5.50) | 6.12<br>(5.86, 6.39) | 0.88<br>(0.56, 1.21) | $9.35 \times 10^{-8}$ | $2.10 \times 10^{-7}$ |
| 23.86 | 5.75<br>(5.46, 6.03) | 6.71<br>(6.42, 7.0) | 0.96<br>(0.59, 1.33) | $4.13 \times 10^{-7}$ | $7.43 \times 10^{-7}$ |

PRS: Polygenic Risk Score. DS: Depressive Symptoms. FDR: False Discovery Rate. Upper and lower 95% confidence intervals for the estimates are given in (parenthesis). The low PRS was for trajectories which were 1 SD below. The high PRS was for trajectories which were 1 SD above. Differences were calculated using the delta method. Analysis were adjusted for sex and the first ten principal components of ancestry.

### Sensitivity Analyses

Negative control analysis using the height PRS demonstrated no evidence for an association with any of the trajectories of depressive symptoms parameters (*P*s ≥ 0.250). Likewise, the PRS for depressive symptoms was not associated with a trajectory for height (*P*s ≥ 0.330) – see Supplementary Tables S17-S18 and Figures S6-S7. Additional sensitivity analysis using a new PRS for major depression found similar, but stronger effects for the trajectories analysis, likely reflecting the increase in power from the new GWAS. However, the overall inferences from the major depression PRS remained the same with the largest difference still occurring between those with higher and lower risk occurring at age 24 with over a one-point difference in the SMFQ (*β*_diff_ = 1.04, 95 CIs = 0.67, 1.42, *P*^FDR^ = 1.24×10^-07^) see Supplementary Tables S19-S20 and Figure S8. One explanation for a stronger observed association between the depressive symptoms PRS and depressive symptoms at age 24, compared to age 10 is measurement variance (i.e., depressive symptoms were measured more accurately at age 24 than at age 10). Tests for measurement invariance between these two ages suggested there was weak measurement invariance between the two ages, with the model taking measurement invariance into account having a better model fit (*x*^2^ = 245.44, *df* = 12, *P* = <.0001). However even when taking measurement variance into account, there was still a stronger association between the PRS and depressive symptoms at age 24 (*β* = 0.105, 95 CIs = 0.06, 0.15, *P* < 0.001), compared to age 10 (*β* = 0.049, 95 CIs = −0.001, 0.098, *P* = 0.051) in the weak invariance model. This suggests that the association between the PRS and depressive symptoms at age 24 reflects a stronger association between the PRS and that depressive symptoms construct, rather than an artefact of measurement variance - see Supplementary Tables S21-S23. Finally, the PRS for depressive symptoms was not associated with the number of occasions an individual completed the SMFQ (*β* = −0.052, 95 CIs = −0.117, 0.012, *P* = 0.114), nor completion of the SMFQ at age 10 but not age 24 (Odds Ratio = 1.053, 95 CIs = 0.996, 1.114, *P* = 0.069). Further information on all analyses are provided in the Supplement.

## Discussion

In this longitudinal cohort study, we focused on a cross-sectional analysis and a growth curve modelling approach that utilised a repeated measures framework to explore the association between genetic risk (as measured by a polygenic risk score for depressive symptoms) and depressive symptoms across adolescence. In the cross-sectional analysis, we found that the PRS was associated with higher levels of depressive symptoms throughout adolescence and early adulthood. There were stronger associations between the PRS and depressive symptoms at older ages that were not explained by measurement invariance. In the growth curve analysis, we found that a higher PRS for depressive symptoms was associated with steeper trajectories of depressive symptoms, that were characterised by greater overall depressive symptoms across adolescence, as well as a greater increase in the rate of change of depressive symptoms.

Our results suggest that a higher PRS is associated with greater depressive symptoms in adolescence and early adulthood and may influence how depressive symptoms change across adolescence and adulthood. Importantly, we see that those with a higher PRS for depressive symptoms begin to have higher symptoms scores between the ages of 12 and 14, suggesting that genetic liability may play a role in the onset of adolescent depression. Such results are consistent with research in high risk family studies [53], and place further emphasis on the idea that depression is not only heritable, but genetic liability may also influence the rate of phenotypic change that is expressed (in this case depressive symptoms across adolescence and young adulthood).

Our repeated measures analysis suggested that the PRS had stronger associations as age increased and that differences in trajectories could in part be the result of varying genetic liability across development. This is consistent with longitudinal twin research showing the genetic contribution to depression increases throughout adolescent development [16, 17]. There are several possible explanations as to why genetic liability may influence change over time. First, genetic liability to depression may act upon biological and hormonal pathways, especially during adolescence [54]. This may result in changes to brain development and hormonal responses that put an individual at greater risk of depression [55]. Second, gene environment correlation with key environmental risk exposures such as stressful life events may increase with age [10, 56, 57]. This in turn may produce indirect pathways to depression that are a result of increased environmental exposures that occur in later development. Third, differences in genetic liability could be the result of measurement error and statistical noise at the varying occasions, but these may be reduced in repeated measures designs [31]. Thus, a repeated measures design that incorporates genetic data may be a more useful model for exploring the temporal association between genetic risk and a developing phenotype.

Previous research has shown that PRS can be included into longitudinal models that examine change in a trait over time [28-30]. Research has suggested that it is also possible to examine genetic influences on age related changes in depression [31]. We were able to expand upon previous work to examine genetic contributions to varying trajectories of depressive symptoms, but specifically in this study we were able to demonstrate how PRS may vary by age. We demonstrated that genetic influences may be age specific (i.e., may begin to onset at different times) which enhances previous research looking at groups of individuals. Our results highlight a useful approach for a repeat measures framework (such as a growth curve model) to quantify the extent to which genetics may influence traits over time (by examining rate of change) which goes over and above traditional cross-sectional research.

This study had several strengths. First, we were able to use a large longitudinal population cohort with repeated assessments of depressive symptoms using the same measure and informant across adolescence to adulthood. Many longitudinal studies with this depth of data change measures or informants, yet we were able to utilise the same measures and informants to characterise trajectories of depressive symptoms across key transitional periods of heightened vulnerability to depression. Second, we were then able to expand upon previous research by using a repeat measures model that is a powerful alternative to cross-sectional analysis. By using the correlation between the repeated measurements, it may be possible to reduce measurement error and boost statistical power, evidenced by the stronger associations at varying ages in the growth curve analysis. Our negative control analysis examining the association between a PRS for height and trajectories of depressive symptoms, and then a depressive symptoms PRS for trajectories of height support this claim and did not show any evidence for non-specific genomic predictors influencing unrelated trajectories. Meanwhile, further analysis using a recent PRS for major depression supported our initial results. Third, we were also able to use the same sample across our analysis as the repeated measures model used full information maximum likelihood (FIML) to account for missing data, which is an advantage of this method compared to the cross-sectional analysis which may require additional methods for missing data.

However, this study had several limitations. First, this study did suffer from attrition as sample sizes varied across ages in the cross-sectional analysis, which may bias our results as missingness is not random. One of the advantages of using the repeat measures model is that we can instead use FIML to account for missing data. This approach also minimises the bias that may be present if each occasion (age wave) represents a different sample. However, even this approach may be biased if the data are missing not at random. For example, genetic risk for depression may predict missing assessments in ALPSAC [24, 58]. This could lead to bias in this study and an underestimation of the true estimate for the trajectories of depressive symptoms and a further underestimation of the genetic contributions to depressive symptoms. We tested this in sensitivity analysis and found that the PRS for depressive symptoms was not associated with the number of times an individual completed the depressive symptoms questionnaire nor rates of attrition. Therefore, this bias is likely to be minimal. Second, depression is a heterogeneous condition and there may be a genetic difference between sum scores, and specific symptoms of depression [59]. Depression symptoms may also differ at different ages [60]. We used the same summary score of depressive symptoms throughout our study, which therefore only captures the sum of depressive symptoms and does not highlight if certain symptoms of depression (i.e., anhedonia, lack of appetite or depressive thoughts) are more related to genetic liability for depressive symptoms. Likewise, tests for measurement invariance indicated that the responses to the depressive symptoms questionnaire may have changed over time, yet the stronger association between the PRS and later occasions was independent of measurement variance. Nevertheless, future research should look to examine how different profiles of depression change across time and how these profiles are independently predicted by genetic risk. Third, our results lack generalisability to all populations as the original GWAS was conducted on individuals of European ancestry, and this may have consequences on further clinical applications [61]. However, research is beginning to capture GWAS of non-European populations [62], and future studies will be able to examine the impact of genetic liability on adolescent depression in other populations. Finally, whilst our results highlight a potential role for PRS in understanding and examining pathways to depression (particularly as PRS are based on genotypes and are at fixed at birth and therefore less susceptible to reverse causation), the amount of variance in depressive symptoms explained by the PRS was low (never more than 1.07%). Whilst this is similar to the effect sizes reported in previous work [19-21, 26], the clinical implications of these results are not clear and PRS should continue to be used for making group-level, rather than individual-level predictions at this stage [63].

In conclusion, we found evidence that a PRS for depressive symptoms associates with measures of depressive symptoms from ages 10 to 24. This PRS was also associated with how depressive symptoms change over time, providing evidence that higher genetic liability to depression is associated with higher trajectories of depressive symptoms (estimated via a higher intercept and slope). Growth curve models that use a repeated measures framework may be a useful tool in genetic analysis, providing greater statistical power/measurement precision and the opportunity to examine changes and variation in depression over time. Our results add to the body of evidence that genetics play a role in the onset and maintenance of adolescent depression and highlight the potential importance of this information for examining pathways to depression.

## Data Availability

Data can be requested from ALSPAC via an online proposal system (www.bris.ac.uk/alspac).

## Acknowledgments

We are extremely grateful to all the families who took part in this study, the midwives for their help in recruiting them, and the whole ALSPAC team, which includes interviewers, computer and laboratory technicians, clerical workers, research scientists, volunteers, managers, receptionists and nurses. Part of this data was collected using REDCap, see the REDCap website for details (https://projectredcap.org/resources/citations/).

